# Navigating sample overlap, winner’s curse and weak instrument bias in Mendelian randomization studies using the UK Biobank

**DOI:** 10.1101/2021.06.28.21259622

**Authors:** Ildar I Sadreev, Benjamin L Elsworth, Ruth E Mitchell, Lavinia Paternoster, Eleanor Sanderson, Neil M Davies, Louise AC Millard, George Davey Smith, Philip C Haycock, Jack Bowden, Tom R Gaunt, Gibran Hemani

**Author notes:** These authors contributed equally (Ildar I Sadreev, Benjamin L Elsworth, Tom R Gaunt, Gibran Hemani).

## Abstract

We performed GWAS on 2514 complex traits from the UK Biobank using a linear mixed model, identifying 40,620 independent significant associations (p<5×10^−8^). We estimate that winner’s curse incurs substantial overestimation of effect sizes in a mean of 35% of discovered associations per trait. We use these results to estimate that the polygenicity of most complex traits is below 10000 common causal variants. We evaluated the impact of winner’s curse on causal effect estimation and hypothesis testing in Mendelian randomization analyses. We show that winner’s curse substantially amplifies the magnitude of weak instrument bias, though any inflation of false discovery rates tends to be low or modest. We designed a process of pseudo-replication within the UK Biobank data to generate GWAS estimates that minimise bias in MR studies using these data. Our resource is integrated into the OpenGWAS platform and enables a convenient framework for researchers to minimise bias or maximise precision of causal effect estimates.

## Introduction

Genome-wide association studies (GWAS) are a powerful tool to identify associations between traits of interest and genetic variants. The rapid growth in genotyped samples, partly due to the emergence of large national biobanks, enables well powered causal inference via Mendelian randomization (MR)^1,2^. MR represents a statistical framework for estimating the causal effect of one phenotype (henceforth the ‘exposure’) on another (henceforth the ‘outcome’), which reduces the potential for biases that often influence observational associations due to factors such as confounding or reverse causation^3^. This is achieved by using results from GWAS to identify genetic variants that are robustly associated with the exposure, to use as instrumental variables. However, there are a number of biases that can affect MR estimates that have been widely documented^4^. In this paper we focus on weak instrument bias in the context of MR performed on GWAS summary level data. Several factors relating to the design of an MR study contribute to the extent to which weak instrument bias manifests, and here we attempt to generate recommendations for addressing this issue in practice along with an extensive GWAS summary data resource based on the UK Biobank.

The problem of weak instruments for MR studies has been described previously^5–8^. MR is predicated on determining the relationship between exposure and outcome through the lens of only the known genetic factors for the exposure, which typically represent a small fraction of the total variance of the phenotype, though this is a fraction of the variance which is less susceptible to confounding and reverse causation. When the core assumptions are met, the difference in the outcome across values of the genotype (instrument) can be ascribed to differences in the exposure. However, if the instrument-exposure association is weak, then a fraction of the observed association is likely to be due to chance associations with confounding factors. The weaker the instrument-exposure association, the larger the fraction of the error term from the SNP-exposure estimation that correlates with confounders. The consequence of this phenomenon is that MR associations obtained using weak instruments will be biased. In the case that the exposure and outcome effects are estimated in a single (i.e. fully overlapping) sample, the bias will be in the direction of the (confounded) observational association. When SNP-exposure and SNP-outcome estimates are obtained from two non-overlapping samples, the bias is in the direction of the null. Partial sample overlap will lead to an estimate that lies between these two extremes. Weak instrument bias is conceptually similar to regression dilution bias: in the single sample setting due to non-differential measurement error and in the two sample setting due to classical measurement error^9,10^. Since the latter bias is always conservative, studies often go to substantial lengths to avoid any sample overlap between the exposure and outcome^8^. However, limiting analyses to studies with no sample overlap discards data, hence reducing statistical power.

Navigating the issue of weak instrument bias in practice requires two further elements that have not previously been considered: the impact of winner’s curse of GWAS estimates on the bias, and whether the bias impacts false discovery rates. Winner’s curse^11^ in the GWAS context describes an ascertainment bias in which the true genetic effect of GWAS ‘significant’ SNPs are on average smaller than the GWAS estimated effect, especially when a strict significance threshold is applied. Despite there being a clear theoretical basis underpinning weak instrument bias, it is tempting to dismiss the issue in the GWAS context because strict significance thresholds are typically employed when selecting instruments. For example, a p-value of 5×10^−8^ relates to an F value of approximately 29, which is much larger than the threshold traditionally used to determine a weak instrument of F > 10. An F-statistic of 10 approximately means that the bias due to weak instruments is no more than 10% of the confounded observational bias in a one-sample setting. This relationship grows in complexity when considering the influence of winner’s curse on the perceived strength of an instrument from GWAS. Under a polygenic architecture of complex traits, a large fraction of effects may be too weak to surpass strict thresholds, but repeated sampling across many true small effects means that some will be overestimated to a sufficient extent that they surpass the threshold. The simplest result of this process is that the effect of genotype on exposure may be overestimated and therefore the causal effect of exposure on outcome in turn underestimated^12^. **Supplementary figure 1** shows the distribution of F-statistics across GWAS in the OpenGWAS database^13^ for the significant associations using the p-value threshold of 5×10^−8^. The mass of associations close to the threshold suggests that there is potential for substantial winner’s curse bias amongst the set of instruments routinely obtained from GWAS and used in MR, and that many apparently strong instruments may in fact be weak.

In this study, we use simulations to demonstrate that weak instrument bias grows dramatically in the presence of winner’s curse, and then explore one, two and three sample study designs for controlling weak instrument bias in two-sample MR. We go on to generate GWAS summary data for 2514 traits in the UK Biobank, and additionally make available split-sample discovery and replication versions of these GWASs for use in MR studies. We use these resources to estimate the extent of winner’s curse in practice, and implications for trait polygenicity. Finally, we explore empirically the impacts of different sample study designs on weak instrument bias and corresponding power and false discovery rates using these GWAS summary datasets.

## Results

### Winner’s curse substantially exacerbates weak instrument bias

Simulations mimicking real GWAS studies were conducted in order to estimate weak instrument bias in MR as a function of sample overlap in the presence of the winner’s curse. We estimated SNP effects on the exposure and outcome using the Inverse Variance Weighting (IVW) according to the schematic diagram shown in **Supplementary Figure 2. Figure 1A** shows the bias for all instruments and for varied magnitudes of confounding. As expected, the direction of the bias is towards null when exposure and outcome samples are independent, and towards the observed (confounded) estimate when there is complete sample overlap. Amongst these simulations the largest bias appears when the set of instruments are weakest and the confounding effect is largest. For example, when there is complete overlap between the discovery and the outcome datasets, our simulated scenario with the weakest instrument and largest confounding effect overestimated the true causal effect by 30%.

**Figure 1:**
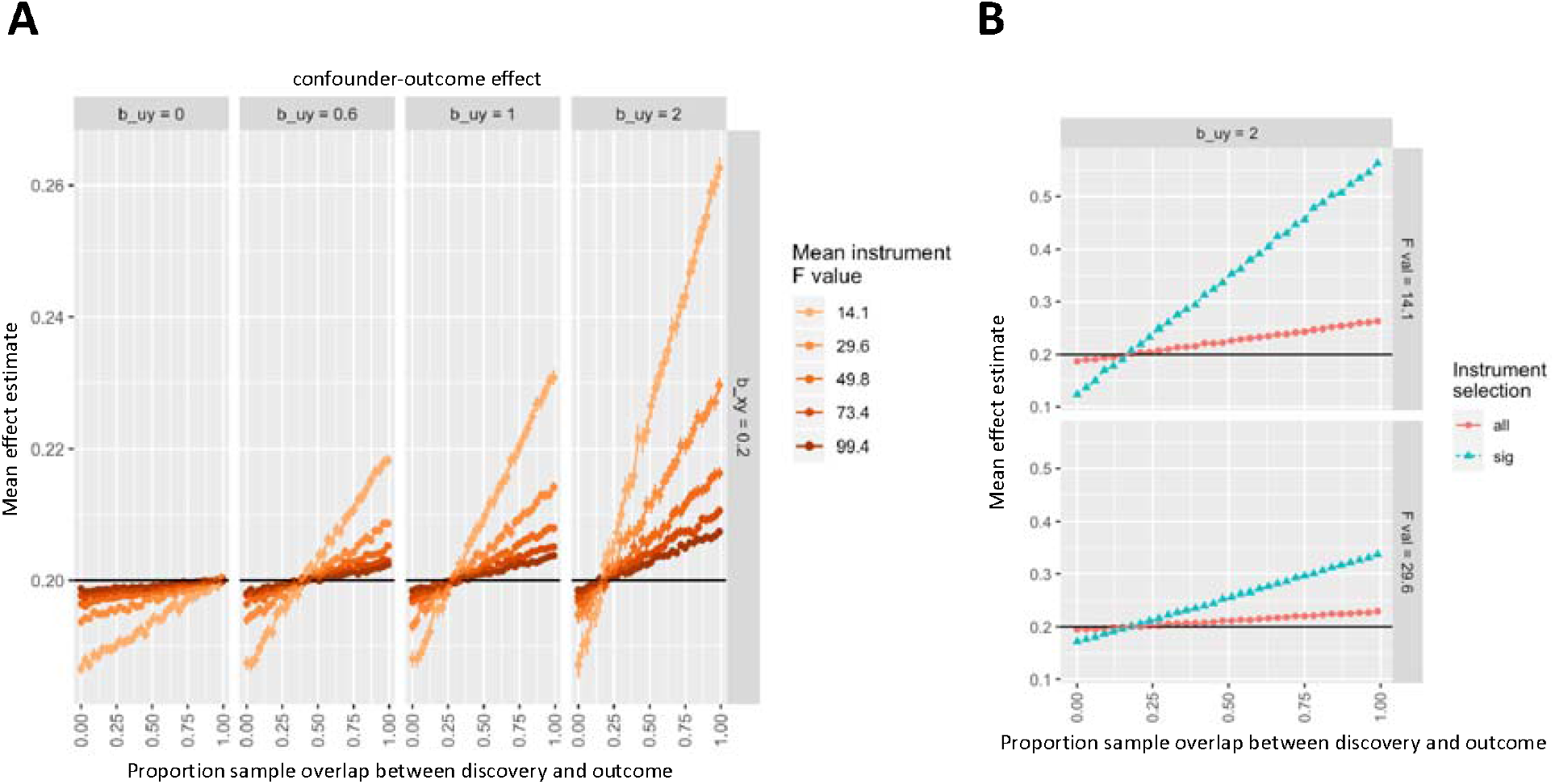
Weak instrument bias, winner’s curse and sample overlap. Solid horizontal line indicates the true simulated causal effect of 0.2 of x on y in arbitrary units. The x-axis for both graphs represents the proportion of the sample overlap between the samples used to estimate the SNP-exposure effect and the SNP-outcome effect. The y-axis represents the mean effect estimate across the simulations for that scenario. A. Simulations depicting weak instrument bias for different effects of confounders on y (b_uy; columns of plots, effects in arbitrary units) and mean instrument strength (F value; coloured lines), in which all simulated instruments were used regardless of SNP-exposure significance level. B. As in A, but looking only at the case of the largest confounding effect and the weakest two instruments. Colours here represent whether all instruments were used (as in A), or only significant associations (p-value of 5×10^−8^) were used, which introduces winner’s curse. Note that difference in scale of the figures in A and B. We observe that the most extreme bias from panel A is substantially magnified by winner’s curse in panel B.

We then expanded the simulations to introduce winner’s curse, where only the SNPs that pass the significance threshold for p < 5×10^−8^ were selected as instruments. **Figure 1B** illustrates that this form of instrument selection dramatically amplifies the weak instrument bias. For example, in our full sample overlap scenario where instruments are weakest and the confounding effect is largest, the percentage bias rises to being 280% larger than the true causal effect. Given conventional GWAS and two-sample MR current practice, it is impossible for weak instruments to not suffer from winner’s curse as the standard significance threshold for instrument selection corresponds to a high F-value.

### Managing bias through choice of study samples

We next used simulations to explore sampling and analytical strategies for overcoming the weak instrument bias issues in the context of a summary-data based MR analysis. We define three potential data sources to generate a summary-MR dataset: the exposure discovery data source D for discovering exposure instruments; an exposure replication data source “R” that shares no sample overlap with “D”, from which discovery effects are re-estimated to overcome winner’s curse; and the outcome data source “Out”, from which the (exposure) instrument effects on the outcome trait are estimated. The samples used to generate GWAS outcome estimates could overlap with the samples used in the discovery and/or replication data sources. We simulated (a) full sample overlap of D and Out, (b) full sample overlap of R and Out, (c) partial sample overlap of both D and R with Out, (d) no sample overlap of either D and R with Out and (e) direct use of D with no replication.

We initially ran the simulations in the unrealistic scenario, where no significance threshold was imposed and no winner’s curse occurred since all the instruments are known. **Figure 2A** and **Supplementary figure 3A** shows that across a range of scenarios the bias in the MR estimate tends to be modest. Next, we introduced winner’s curse into this analysis by applying a significance threshold for instrument selection. **Figure 2B** and **Supplementary figure 3B** demonstrates that the level of bias in the MR estimate was dependent upon the sampling strategy and the manner in which winner’s curse was handled. The theoretically ideal scenario is where winner’s curse is eliminated by re-estimating the discovered instrument-exposure effects in an independent sample, and the SNP-outcome estimates come from a third independent sample (scenario d above)^14^. Our simulations confirm that for this scenario the MR estimators are unbiased. A strategy that is used commonly in practice is to use discovery SNP-exposure effect estimates without replication, and to allow sample overlap between the discovery and outcome data sources. In this scenario the bias of the MR estimate occurs in the direction of the observational association and is substantially magnified in comparison to the case where there is no winner’s curse. This bias in MR only occurs if the observational association is biased by confounding. Correspondingly, when there is no overlap between the discovery and outcome data source samples, the bias is large but in the direction of the null hypothesis.

**Figure 2:**
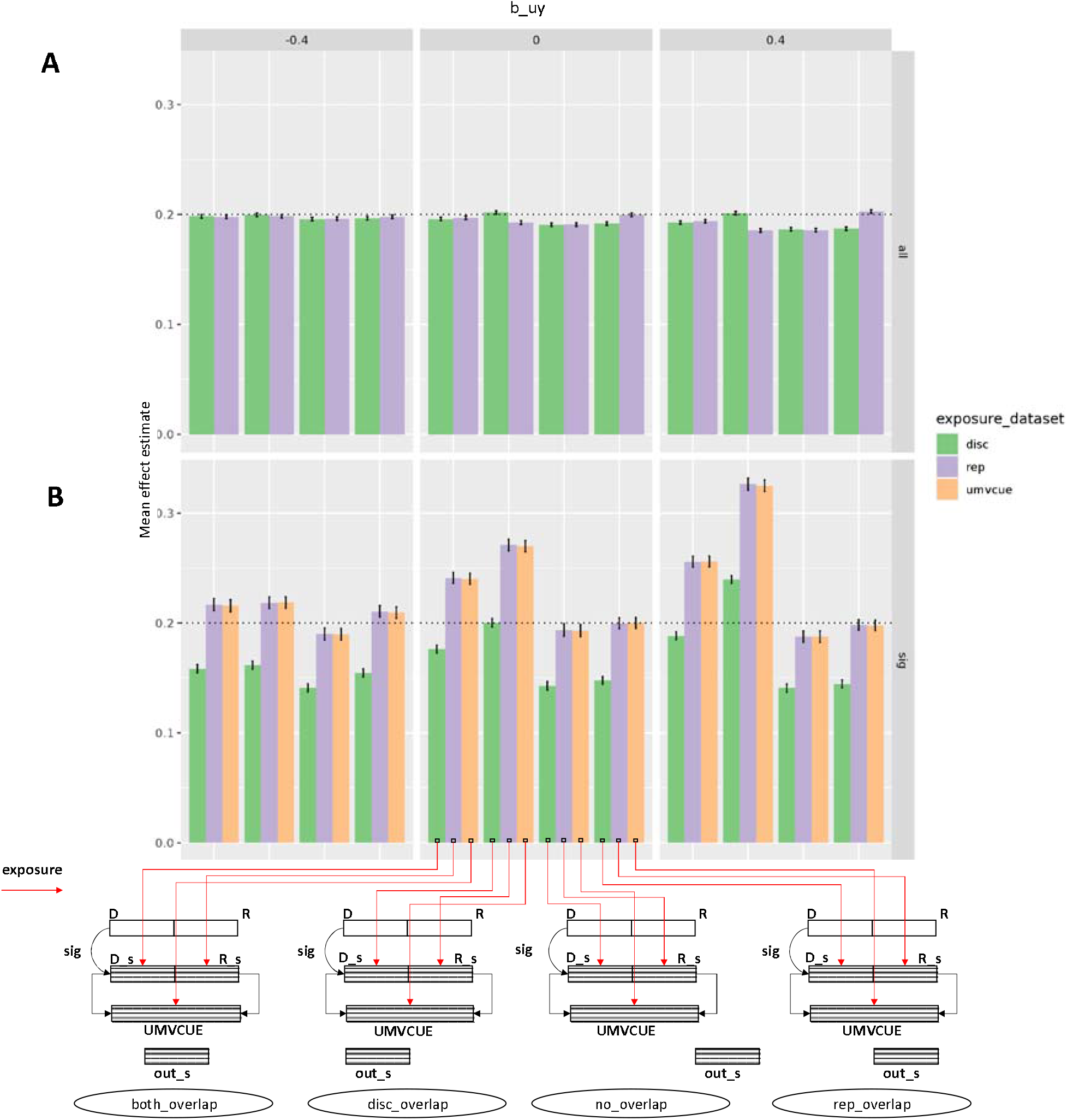
Performance across different sample configurations. We extended the simulations in Figure 1 to allow the use of a replication dataset to re-estimate the SNP-exposure effects, thus avoiding winner’s curse. Panel A represents the scenario where all instruments were used regardless of significance level. Panel B is identical to panel A except the simulations only used instruments that were significantly associated with the exposure, thus introducing winner’s curse. The true causal effect of X on Y is 0.2 (arbitrary units). Columns of plots relate to three different magnitudes of confounding, with effects in arbitrary units on X and Y. The y-axis represents the mean effect estimate across the simulations for each scenario. Sets of bars across the x-axis represent the degree of sample overlap between the SNP-outcome dataset and the SNP-exposure dataset. Colours of bars represent the source of the samples used for the estimate of the SNP-exposure association. The sample configuration schematic shows the sample origin of the SNP-exposure and SNP-outcome effect estimates for each bar. Here, D = discovery dataset (for identifying SNPs as instruments); R = replication dataset (for estimating SNP-exposure effects); UMVCUE = Estimates obtained by combining the discovery and replication exposure estimates using the Uniformly Minimum Variance Conditionally Unbiased Estimator; and out_s = outcome dataset (for estimating SNP-outcome effects).

The pattern of bias changes when winner’s curse correction is introduced through the use of independent replication to re-estimate the SNP-exposure effects. In the case that the SNP-outcome association is estimated using samples with full overlap with the original instrument discovery data source (scenario a), bias in the MR estimate persists. However, in the case that the SNP-outcome association is estimated in a data source with samples overlapping with the replication data source (scenario b), bias of the MR estimates is minimised to be very similar to the three sample scenario despite requiring only two samples. In addition to using the raw replication estimates, we can also attempt to improve their precision by combining with the discovery associations using the Uniformly Minimum Variance Conditionally Unbiased Estimator (UMVCUE)^15,16^. We note that either approach provides comparable results.

An issue pertaining to weak instrument bias which is seldom investigated is the impact on hypothesis testing, and in particular, whether the false discovery rate is elevated due to the bias. **Figure 3** demonstrates that across all scenarios the false discovery rate is relatively constant, despite the large disparities in the level of bias, though. The two sample scenario in which outcome samples overlap with the replication (scenario b) approximates the ideal three sample scenario in terms of hypothesis testing (scenario e). Performing this two sample scenario in both directions (e.g. alternating data sources A and B for the exposure and outcome), and meta-analysing the results, gives rise to substantially improved power without increasing false discovery rates.

**Figure 3:**
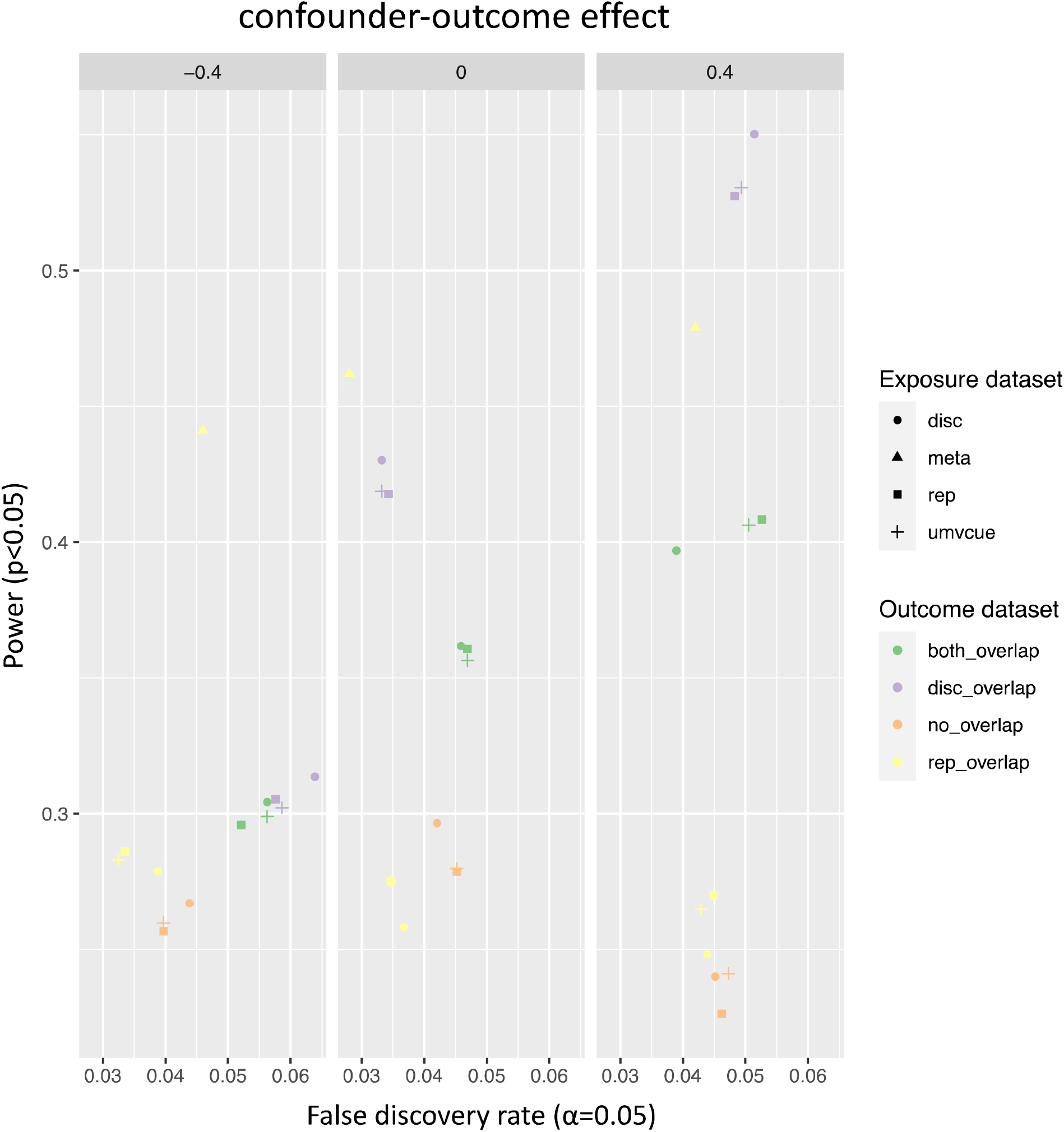
Impact of sample configurations on hypothesis testing. Using the same simulation data for significant SNPs from Figure 2B, the false discovery rate for null associations (x-axis) is plotted against the power of non-null associations (y-axis). Point shapes depict the source of the SNP-exposure effect estimates, and point colours represent the source of the SNP-outcome effect estimates. The outcome dataset names correspond to the ones schematically illustrated in Figure 2B. The two sample scenario in which outcome samples overlap with the replication (yellow squares) approximates the ideal three sample scenario (orange squares). Performing this two sample scenario in both directions (e.g. alternating datasets A and B for the exposure and outcome), and meta-analysing the results (yellow triangles) results in substantially improved power without increasing false discovery rates.

In summary, winner’s curse magnifies weak instrument bias. Adjusting for winner’s curse with an independent replication dataset does not solve the weak instrument problem when there is substantial sample overlap between the discovery and the outcome samples. Any overlap between replication and outcome samples has a small impact on bias, and the impact on the false discovery rate of most weak instrument bias scenarios appears to be minimal.

### A resource of GWAS summary data from the UK Biobank

We performed GWAS on 2514 heritable traits measured in the UK biobank using a linear mixed model on 460k European samples. After strict quality control we identified at least one significant genetic association (p < 5×10^−8^) for 1131 traits, and summing across all traits 40,612 independent (clumping R^2^ < 0.001) associations overall, involving 22,181 unique variants.

Given that a major use of these summary statistics will be for MR analyses, in which the exposure and outcome estimates could be obtained from amongst these results, our simulations indicate that the resource suffers from two limitations. First, MR estimates amongst these UKB GWAS will suffer from both winner’s curse and sample overlap, meaning that the weak instrument bias in the direction of the observational association will be magnified. Second, if a dataset from this resource is analysed (e.g. as the exposure) with an outcome dataset obtained from non-overlapping samples, the winner’s curse will magnify any bias that arises in the direction of the null. Therefore we re-generated GWAS summary data for these traits, dividing the samples into two sets and generating two new sets of summary statistics for each trait. These data enable MR analyses resembling scenario b described above.

### Optimising discovery/replication sample splits for MR analysis

A number of factors need to be considered in determining the proportion by which to split the samples. We conducted simulations that showed that the optimal split depends on a) whether power or bias are being optimised and b) the genetic architecture of the traits. For traits with a high heritability, our simulations showed that the power of an MR analysis was maximised when the exposure discovery sample was smaller than the exposure replication sample (**Figure 4A**), though this relationship was reversed for traits with a low heritability and was conditional on at least one instrument being discovered. Weak instrument bias generally reduced as the exposure replication sample grew larger (**Figure 4B**). By contrast, the number of instruments discovered increased almost linearly as the fraction of the sample used for the exposure discovery grew larger (**Figure 4C**). Selecting a single splitting proportion across all traits is practically most advantageous, because then any combination of traits can guarantee complete sample independence between the discovery and outcome datasets. Hence we determined that splitting the UK Biobank sample into two sets of 50% each would provide the most flexibility and our simulations indicate that this does not incur major statistical disadvantages. Researchers performing specific analyses may wish to determine the sample split based on the heritabilities of the traits under their consideration.

**Figure 4:**
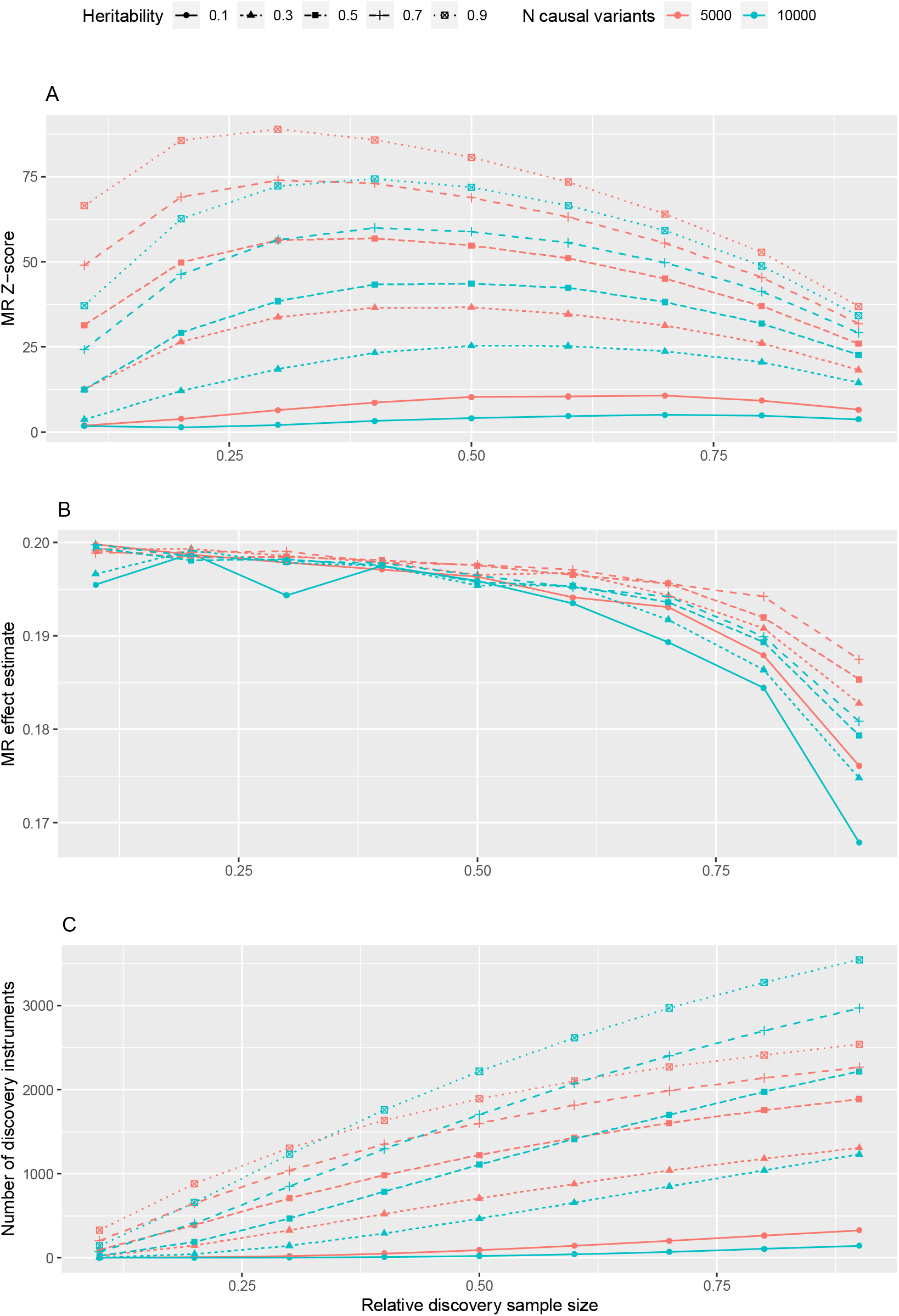
Sample split simulations. The relationship between SNP-exposure discovery and replication sample sizes on the performance of MR analysis. The x-axis represents the fraction of the sample used for instrument discovery. The y-axis in panel A represents the Z-score for non-null simulations (i.e. the power); in panel B the MR effect estimate, where the true causal effect is 0.2; and in panel C the number of discovery instruments. The number of simulated causal variants is depicted by point and line colours, the heritability of the trait depicted by point and line shapes.

### Analysis of winner’s curse in the UK Biobank

We split the 460k European samples into two equally sized random subsamples (A and B), and re-analysed each of the 2514 UKBB traits within each subsample. We found at least one significant genetic association (p < 5×10^−8^) in either the A or B subsample for 590 traits. There was a median of 3 significant associations amongst traits with at least one association. In total 13,673 and 13,723 independent associations were discovered in subsamples A and B, respectively. Amongst these instruments we identified 681 as being ‘weak’, defined as having an association at p < 5×10^−8^ in one subsample but having a replication F value < 10 in the other subsample. However, 2589 instruments had substantially overestimated effect estimates in which the replication estimate was lower than the 2.5% confidence interval of the discovery estimate. By this definition, 14% of traits had only over-estimated genetic associations. The mean proportion of instruments being substantially overestimates per trait was 35% (median=23%).

We next conducted simulations to contrast our empirical estimates of winner’s curse against expectations for a range of genetic architectures and sample sizes. Under polygenic architectures, our results indicate that the fraction of detected variants that are weak or overestimated decreases as the number of discovered variants increase (**Figure 5A**), suggesting that the problem of weak instrument bias will likely reduce as GWAS sample sizes continue to grow because the fraction of instruments that are substantially overestimated due to winner’s curse will shrink.

**Figure 5:**
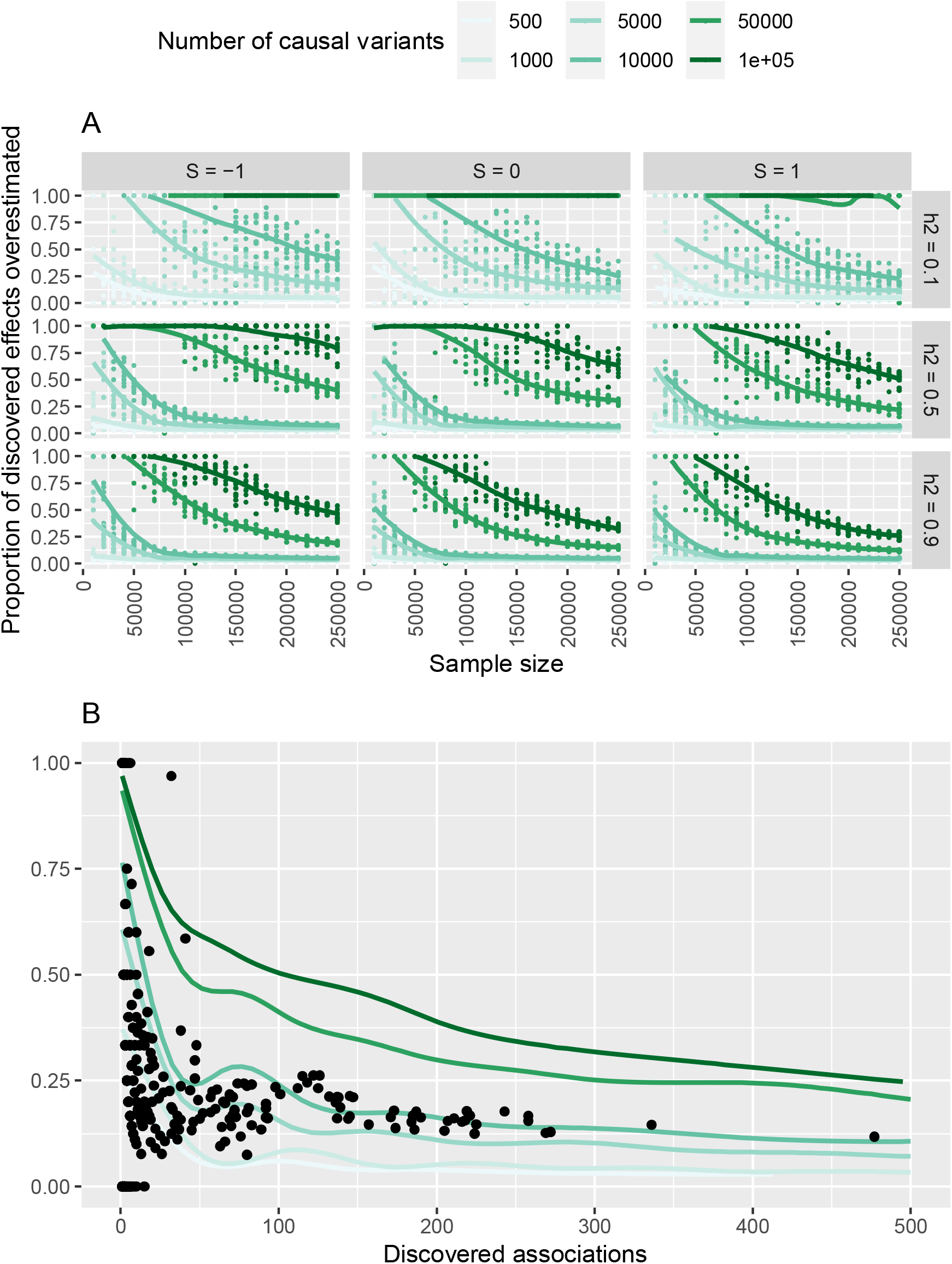
The extent of winner’s curse in complex trait GWAS. Panel A shows the fraction of discovered effect sizes that are substantially overestimated on the y-axis (the 2.5% lower bound of the absolute effect estimate is larger than the true effect size), against sample size (x-axis). Rows of plots correspond to trait heritability (h2), columns of plots correspond to a coefficient representing natural selection (S), and colours represent the polygenicity. As selection gets more positive, heritability gets lower, and sample size gets smaller, the fraction of discovery effect estimates that are overestimated grows larger. Given sufficient power the fraction of discovery SNPs that are substantially overestimated becomes very small. Panel B takes the simulated datasets used to generate panel A and superimposes the results from GWAS discovery and replication analysis of 2514 traits (black points), showing that they generally follow a pattern of polygenicity involving 10000 causal variants or fewer.

Next we used these observations to infer the polygenicity of the analysed traits, by superimposing the UK Biobank results against a range of simulation scenarios. We found that the observed pattern of winner’s curse in the empirical data for the majority of traits is consistent with scenarios in which fewer than 10000 causal variants contribute to the trait (**Figure 5B**).

### Empirical analysis of weak instrument bias in the UK Biobank

To examine the extent to which weak instrument bias, winner’s curse and sample overlap influence MR results empirically, we performed pairwise MR analysis of all 2514 traits across a range of different sample configurations using the GWAS summary data generated for these traits. Results described here are based on the IVW method, but similar trends are seen for other summary-based MR methods such as the weighted median, weighted mode or Egger regression. We conducted the MR analyses in the sample configurations described in **Table 1**.

**Table 1:** Sample configurations used for empirical MR analyses using GWAS summary datasets from the UK Biobank.

| SAMPLE<br>CONFIGURATION | INSTRUMENT<br>DISCOVERY | SNP-EXPOSURE<br>REPLICATION | SNP-OUTCOME<br>EFFECT<br>ESTIMATE | NOTES |
| --- | --- | --- | --- | --- |
| 0 | Full GWAS dataset | None | Full GWAS dataset | Winner's curse, complete sample overlap |
| 1A | Subsample A | Subsample B | Subsample B | No winner's curse, outcome sample overlaps only with replication |
| 2A | Subsample A | None | Subsample B | Winner's curse, no sample overlap |
| 3A | Subsample A | Subsample B | Subsample A | No winner's curse, outcome sample overlaps with discovery |
| 4A | Subsample A | None | Subsample A | Winner's curse, complete sample overlap |
| 1B | Subsample B | Subsample A | Subsample A | No winner's curse, outcome sample overlaps only with replication |
| 2B | Subsample B | None | Subsample B | Winner's curse, no sample overlap |
| 3B | Subsample B | Subsample A | Subsample B | No winner's curse, outcome sample overlaps with discovery |
| 4B | Subsample B | None | Subsample B | Winner's curse, complete sample overlap |

**Table 2:** Summary of MR analysis for age at death on duration of walks for various sample configurations.

| SAMPLE CONFIGURATION | INSTRUMENT DISCOVERY | SNP-EXPOSURE REPLICATION | SNP-OUTCOME EFFECT ESTIMATE | NOTES | MR for duration of walks on age at death |  |  |  |
| --- | --- | --- | --- | --- | --- | --- | --- | --- |
|  |  |  |  |  | beta | se | p-value | nsnp |
| 0 | Full GWAS dataset | None | Full GWAS dataset | Winner's curse, complete sample overlap | 1.1 | 0.33 | 0.00085 | 13 |
| 1A | Subsample A | Subsample B | Subsample B | No winner's curse, outcome sample overlaps only with replication | 5.3 | 1.50 | 0.00046 | 2 |
| 2A | Subsample A | None | Subsample B | Winner's curse, no sample overlap | 2.4 | 0.69 | 0.00044 | 2 |
| 3A | Subsample A | Subsample B | Subsample A | No winner's curse, outcome sample overlaps with discovery | 2.2 | 1.51 | 0.14506 | 2 |
| 4A | Subsample A | None | Subsample A | Winner's curse, complete sample overlap | 1.0 | 0.69 | 0.14555 | 2 |

We use configuration 1A/1B, which has no winner’s curse and no discovery-outcome sample overlap, as the baseline in the comparisons that follow. We observed that introducing only winner’s curse (configuration 2A/2B) or only sample overlap between the instrument discovery and outcome datasets (configuration 3A/3B) induced a moderate bias (Pearson correlation of effect sizes versus configuration (1A/1B) of r = 0.79 and 0.76, respectively). However, the joint introduction of winner’s curse and sample overlap (configuration 4A/4B) led to a more magnified bias (r = 0.43) (**Figure 6**). Most traits analysed here contain a mixture of stronger and weaker instruments, with most instruments being strong. To check if the results also hold when all the instruments are weaker, we performed the comparisons against configuration 1A/1B again but using MR analyses with only weak instruments (**Supplementary figure 4**). We found that the bias is exacerbated when only weaker instruments are used (e.g. the joint introduction of winner’s curse and sample overlap (configuration 4A/4B) further magnified the bias, r = 0.1).

**Figure 6:**
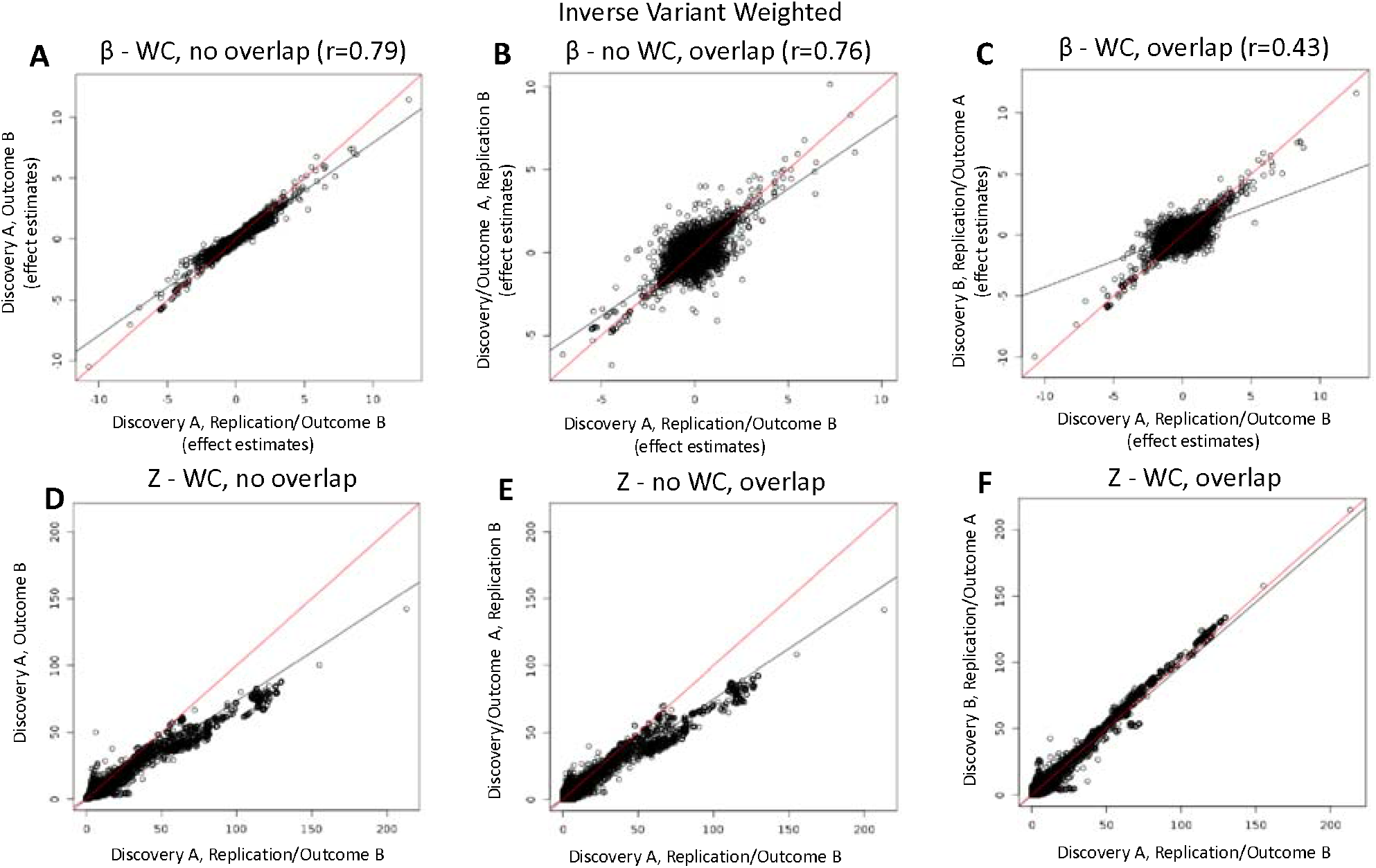
Empirical MR analyses compared across different sample configurations. A-C show the effect estimates of the “no winner’s curse, no sample overlap” scenario (configuration 1 from Table 1) on the x-axis against the configurations 2, 3, and 4 on the y-axis, respectively. Similarly, panels D-E represent the correspondence of Z-scores of configuration 1 on the x-axis against configurations 2, 3, and 4, respectively. Scenario 1 – replication used as an exposure, no overlap between discovery and outcome. Scenario 2 – discovery used as an exposure, no overlap between discovery and outcome. Scenario 3 – replication used as an exposure, full overlap between discovery and outcome. Scenario 4 – discovery used as an exposure, full overlap between discovery and outcome.

By contrast, the test statistics were much more strongly related across the different scenarios (including when only weak instruments were used). Of the associations in configuration 1A/1B with p > 0.05, only 5.8% were found to have p < 0.05 in configuration 4A/4B, where there is both winner’s curse and discovery-outcome sample overlap, suggesting a minimal increase in FDR. Similarly, of the associations in configuration 1A/1B with p-values < 0.05, only 6.5% of those in configuration 4A/4B returned p > 0.05, indicating relatively minimal impact on power.

Finally, we looked at the traits in UK Biobank that could likely give a high winner’s curse bias and performed the analysis in the three following ways: a. full MR, b. using only half of the randomly split samples for the discovery and outcome (corresponds to both sample overlap and winner’s curse) and c. using the replication dataset that overlaps with the outcome (no winner’s curse, sample overlap only with the replication). The results of MR for causal effect of duration of walks on age at death are summarised in **Table 1**. Our analysis suggests that applying the approach proposed in this paper of using the replication and the outcome from the same dataset with no overlap with the discovery dataset gives a different result compared with full MR and the sample overlap and winner’s curse scenarios. In particular, the proposed method gives higher association estimate as well as a significant p-value, whereas the worst case scenario of the full sample overlap and winner’s curse leads to a smaller estimate and a non-significant p-value.

## Discussion

MR has proven to be a powerful tool for causal inference^17,18^. However, uncertainty remains around the best approaches to minimise bias due to weak instruments. In particular, the impact of weak instrument bias on MR results in the presence of winner’s curse has been neglected, as has the relevance of weak instrument bias to the performance of hypothesis testing. We have examined these using both simulations and empirical analysis, and generated a resource of GWAS summary data that offers flexibility in analysis. Much attention is given to methodological advances that attempt to minimise bias in MR analysis, however it is worth pausing to consider what features of the approach are of most practical importance. In practice, for binary outcomes the magnitude of the effect size often matters less than being able to correctly declare the presence or absence of a causal relationship and its orientation (i.e. be valid under the null hypothesis). In this study, we have illustrated that weak instrument bias can be considerably larger than has previously been thought, due to the presence of winner’s curse enabling weak instruments to appear to be much stronger than they truly are.

Weak instrument bias has been conceptualised to take place due to the sampling error in the variance estimated in the exposure by the instrument. In the case that there is sample overlap, this sampling error will be correlated with confounders, thus biasing the two-sample MR estimate in the direction of the observational association. In the case of no sample overlap it will represent random noise, thus biasing the two-sample MR estimate in the direction of the null. One explanation for why winner’s curse magnifies weak instrument bias is that significant associations are ascertained to have larger sampling variance (i.e. due to smaller effect sizes or lower minor allele frequency), and therefore the fraction of the variance that is correlated with confounders (in cases where there is sample overlap) or random noise (in cases where there is no sample overlap) will be maximised.

We have developed GWAS summary data from different sample configurations that trade off the impact of weak instrument bias for statistical power, giving flexibility for analysts to determine what is of most relevance to their study. It also provides a convenient framework for sensitivity analyses of the potential impact of weak instrument bias. Analyses can be performed using the summary data in a one-sample setting which will maximise the statistical power (due to larger discovery sample sizes finding more instruments, and larger outcome sample sizes improving the precision of the MR estimate), though the combined impacts of winner’s curse, weak instrument bias and sample overlap will likely magnify the bias in the direction of the observational association in this case. We have provided a resource of genetic associations in subsamples in A and B of the UK Biobank which can be used for analysis of traits specifically within the UK Biobank, which is likely to substantially reduce bias as long as the outcome dataset is independent of the discovery dataset. Alternatively, subsamples A and B can be used to define unbiased SNP-exposure effect estimates to be used in a three-sample MR analysis, where an outcome trait is obtained from an independent set of samples. Another possibility that might be promising, is to meta-analyse two scenarios from **Table 1**, namely 1A and 1B, which correspond to the scenarios where there is a full overlap between the exposure replication and outcome datasets, in two independent configurations, respectively. Our simulations suggest an increase in power for this case (**Figure 3**).

Extant GWAS summary data likely harbour relatively large fractions of effect estimates that are substantially overestimated, though we predict that future GWAS will return a diminishing impact of winner’s curse in effect estimate bias as sample sizes continue to grow.

Our simulations show that in a polygenic model, as the number of detected variants in a GWAS increases, the fraction that are substantially overestimated reduces (**Figure 5**). In particular, we observe a the contribution of weak instruments to GWAS significant associations to approach zero in GWASs with sufficient statistical power to detect thousands of independent variants. Therefore, dividing future large biobanks into discovery and replication datasets may not be warranted if the impact of winner’s curse is low. However, a resurgence in the interests of using family data to avoid demographic and familial structural biases in GWAS does arrest the progress made in improving power with larger population-based analyses. For these smaller studies, winner’s curse and weak instrument bias will remain of relevance for a longer period of time. While weak instrument bias might reduce as sample sizes increase, discovery of more instruments may introduce other biases relating to mechanisms such as horizontal pleiotropy or trait-driven selection bias.

An alternative approach could be combining multiple SNPs into a single instrument and use this multi SNP instrument for the analysis. However, the IVW and PRS estimates are largely equal, suggesting they are likely to be similarly impacted by the winner’s curse. Performing a one-sample MR analysis using CUE and SNPs selected from an external sample could help^7^. In this case, discovery SNPs would be taken from an external GWAS study and used in an individual-level one-sample MR analysis in UK Biobank. However, this suggests that one needs to have an additional external GWAS source as well as access to the individual-level UKBB data. To note, the GWAS summary results we generated in this work, freely available in OpenGWAS, enables other to conduct MR analyses using our improved GWAS estimates with summary data alone.

A number of limitations exist in the current study. We have not evaluated the extent of winner’s curse for the complete sample GWASs because of a lack of external data to re-estimate discovery effect sizes, though our simulations indicate that the sample size will need to increase more before bias due to winner’s curse plateaus. Rather than performing GWAS of all 2514 traits on subsample A and subsample B, we only re-estimated the 40,612 complete sample discovery associations in the A and B subsamples in order to reduce computational burden. The consequence of this is that we will not identify any particularly weak instruments that were chance significant in half the sample size and not in the full sample. The GWAS analyses were performed on disease traits which have elevated false discovery rates in BOLT-LMM when case numbers are low. While other methods have emerged that address this problem, we have instead aggressively filtered out any variants that have a minor allele count < 90, and have only analysed traits with at least 1000 cases (**Supplementary Note 1**). We conducted our weak instrument bias simulations to explore the extent to which winner’s curse affects the magnitude of bias. However, our simulations depicted simple traits with relatively small numbers of variants, and they do not reveal what the relative bias would be expected for large scale GWAS on polygenic traits as this would be computationally prohibitive. Instead, we conducted empirical analyses to compare the difference in effects and hypothesis testing results across different sampling strategies. In these analyses we do not have knowledge of the truth, and so have limited our conclusions to the context of magnitude of difference relative to the best available sample configuration. Finally, we observed that UMVCUE made minimal improvements in analysis over solely using the replication dataset to obtain unbiased instrument effect estimates. This is most likely due to the discovery/replication split being 50%, and the value of UMVCUE is likely to be greater when the replication sample is smaller^15,16^.

Overall, we have shown that weak instrument bias is substantially magnified by winner’s curse, but is unlikely to introduce major problems with hypothesis testing. We have made available GWAS summary data on 2514 traits with different sample configurations through OpenGWAS. We recommend using the full sample data to maximise power in main analyses and using the split sample data for sensitivity analyses, to evaluate whether the interpretation of the results might change when reducing bias from the winner’s curse.

## Methods

### Weak instrument, winner’s curse and sample overlap simulations

Simulations were conducted to evaluate the extent to which instrument strength attenuated the bias in MR due to sample overlap. Simulation parameters were identical to those in Burgess et al (2016)^8^ except we also included larger SNP effect sizes that will produce F statistics that are more in line with those seen in GWAS. Simulations were conducted in two ways:

1. 20 instruments with the same simulated effect size were all used to obtain the IVW estimate (all)
2. Only those instruments among the 20 simulated that had p < 5×10^−8^ were used to obtain IVW estimate (sig), to mimic the standard procedure used in GWAS

The simulations were conducted with different confounder effects and with causal effects of x on y 0.2. The simulations included r^2^_gx_ ∈ {0.04,0.08,…,0.24} meaning that some simulations had all weak instruments weak and the others all strong. We simulated x as

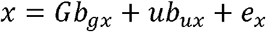

Where G is the matrix of genotypes, standardised to column means of zero and variance 1,*b*_*gx*_ is a vector effects whereby each element j is

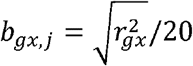

The confounder *u* ∼*N*(0,1) has effect 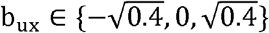 and 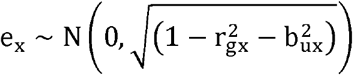. We constructed y to be a function of causal influences from x and u as

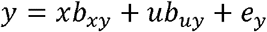

where 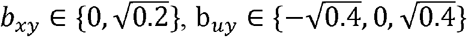, and 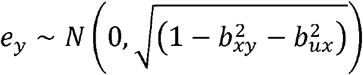.

We varied the proportion of samples overlapping to estimate the 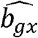 and 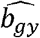 associations ranging from 0 to 100% overlap, and used the association estimates to obtain estimates of 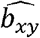 using the inverse variance weighted (IVW) meta-analysis of Wald ratios when multiple instruments were significant at p < 5×10^−8^ (or all were being used regardless of significance), or the Wald ratio if only a single instrument was significant. Each scenario was repeated 10000 times. The simulations used the simulateGP R package https://github.com/explodecomputer/simulateGP/.

### Sampling strategy simulations

We next conducted similar simulations but included the availability of an independent replication dataset for the SNP-exposure estimates. The parameters were identical to those used in the previous simulations except we only used a value of 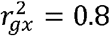. We simulated 3000 individuals in the discovery sample for the SNP-exposure estimates, 3000 independent samples in the replication sample for the SNP-exposure estimates, and 3000 individuals for the SNP-outcome associations. The SNP outcome samples were either drawn from an independent sample, the same sample as the exposure discovery, the same sample as the exposure replication, or an equal mixture of the exposure discovery and replication samples. The SNP-exposure estimates were taken either from the discovery sample, the replication sample, or by combining them using the uniformly minimum variance conditionally unbiased estimator (UMVCUE)^16^.

### Complete genome-wide associate studies

Genotype data from UK Biobank^19^ were quality controlled according to the protocol outlined in^20^. Briefly, this involved removing individuals who did not genetically cluster in the European subsample using an in house k-means clustering algorithm and removing genetic variants that had low minor allele frequencies and imputation quality scores. The final dataset contained 463,005 individuals (this number excludes standard exclusions such as sex mismatch, sex chromosome aneuploidy, outliers in heterozygosity and missingness rates – 1812 individuals but at the same time includes related individuals) and 12,370,749 SNPs. We initially identified approximately 20000 binary, continuous or ordinal traits using the PHEASANT parsing pipeline. We then filtered this set to remove any disease traits that had fewer than 1000 cases amongst the retained individuals, resulting in 2514 traits. GWAS was performed for each trait separately using the BOLT-LMM (version 2.3.2) software. We included 143,006 model SNPs to capture the polygenic background effect and performed strict LD clumping (window size = 10Mb and LD r2 = 0.001) on the resultant GWAS summary statistics to obtain a set of independent instruments for subsequent MR analysis. Sex and genotyping array were included as covariates. The GWAS analyses were performed on disease traits which have elevated false discovery rates in BOLT-LMM when case numbers are low. While other methods have emerged that address this problem, we have instead aggressively filtered out any variants that have a minor allele count < 90.

### Sample split simulations

We conducted simulations using a three-sample MR scenario in which there is a SNP-exposure discovery dataset, SNP-exposure replication dataset, and SNP-outcome dataset, each with non-overlapping samples. For computational efficiency we simulated summary statistics directly for *M* ∈ {5000, 10000} causal variants. The SNP-exposure effects were sampled as

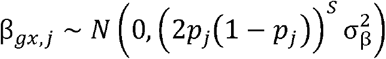

where 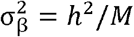, assuming that the variance of the exposure is 1 and S is the selection coefficient. To obtain the underlying *β*_*gy,j*_ effects we multiplied the SNP-exposure effects by the causal effect β_*xy*_= 0.2 for all simulations. The expected standard errors for each effect were calculated as

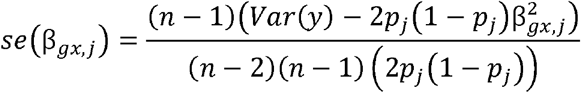

again setting var(y) = 1 for all simulations, and where *p*_*j*_ is the allele frequency of SNP j sampled from a uniform distribution. We then sampled effect estimates for each of the three samples using

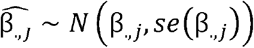

We split a total sample size of 450000 into discovery and replication datasets (ranging from 10% discovery to 90% discovery), and used the same sample size for the outcome dataset as the replication dataset (though with independent draws implying no sample overlap). We fixed S to be 0 in these simulations. Discovery SNP-exposure associations were defined as those with p < 5×10^−8^ and replicated by looking up the association in the replication SNP-exposure dataset. MR analysis was conducted using only instruments that were significant in the discovery but using the replication effect estimates. Causal effect estimates were obtained using the Wald ratio method if only one significant SNP was available, or the IVW method if more than one was available.

### Discovery/replication genetic analyses

To generate a resource for MR analyses that minimises the impact of winner’s curse and sample overlap, we set out to generate a set of GWAS summary statistics for each trait from two independent samples. The entire UK Biobank dataset that was used to generate the full GWASs was randomly split into subsample A and subsample B of equal sizes (the same samples for each trait). We then took the set of 22,181 independent full GWAS discovery SNPs and generated summary data for these SNPs only using BOLT-LMM, and using the same parameters and background SNPs as in the full GWAS analysis. We considered SNPs to be instruments for a trait in subsample A (B) if they had previously been identified as an independent hit in the full GWAS for that trait, and if they had a p-value < 5×10^−8^ in the new subsample A (B) analysis. This means that we only discover SNPs in the subsample that would have been discovered in a full GWAS, but miss SNPs that might have been discovered in the subsample that were absent in the full sample. This latter set of SNPs is likely to be extremely small, and represent very weak instruments with large winner’s curse.

### Winner’s curse simulations

We conducted simulations to estimate the fraction of discovery instruments that are likely to be overestimated or weak across a range of scenarios. The simulations were conducted in the same manner as the sample split simulations, but generating only 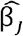 discovery summary statistics and comparing them against the true β_*j*_ effects. Having simulated M causal variants, and selecting only those that were significant at p < 5×10^−8^, we then evaluated if the absolute 2.5% lower confidence value was larger than the true effect size (denoting a substantially overestimated effect), or if the expected effect F value under no ascertainment would have been below 10 (denoting an ascertained weak instrument). We evaluated the following scenarios:

- Sample size *N* ∈ {10000, 20000, …, 500000}
- Number of causal variants *M* ∈ {500, 1000, 5000, 10000, 50000, 100000}
- *h*^2^ ∈ {0.1,0.5,0.9}
- *S* ∈ {-1,0,1}

And repeated each scenario 10 times.

### Mendelian randomization (MR)

For each trait that had an instrument in either the full sample, the A subsample or the B subsample, we performed MR analysis of that as the exposure against all other 2514 traits. In some cases traits had instruments in the full sample analysis but not in either of the subsamples, meaning no comparison could be made. When instruments were available we conducted MR analyses in the sample configurations described in **Table 1**. Where a SNP-exposure replication dataset was used, the instrument discovery dataset was used to select instruments and the SNP-exposure replication dataset was used to estimate the effect sizes, otherwise the discovery effect sizes were used for the SNP-exposure estimate. Two-sample MR was performed in R version 4.0.2 using the R package “TwoSampleMR” version 0.5.5 and the function “mr()”. The methods used for this analysis included Wald ratio when only one instrument was available, and primarily the Inverse variance weighted method when multiple instruments were available. Additional analyses were conducted with the weighted median and weighted mode estimators for cases when three or more instruments were available. The parameters used for the MR methods remained default.

## Data Availability

The GWAS summary data generated as part of this study are available in the OpenGWAS database under batch IDs ukb-b, ukb-ba, ukb-bb at https://gwas.mrcieu.ac.uk/.
Code used to generate the GWAS summary data is available here: https://github.com/MRCIEU/BiobankGWAS/
Code used for the other statistical analysis in the study is available here: https://github.com/isadreev/UKBB_replication

## Acknowledgements

This research was funded in part by the Wellcome Trust and the Royal Society [208806/Z/17/Z] and the UK Medical Research Council (MRC Integrative Epidemiology Unit MC_UU_00011/1 and MC_UU_00011/4). NMD is supported by a Norwegian Research Council Grant number 295989. JB is supported by an Expanding Excellence in England (E3) research grant awarded to the University of Exeter. For the purpose of Open Access, the author has applied a CC BY public copyright licence to any Author Accepted Manuscript version arising from this submission.

## Data availability

The GWAS summary data generated as part of this study are available in the OpenGWAS database under batch IDs ukb-b, ukb-ba, ukb-bb at https://gwas.mrcieu.ac.uk/.

Code used to generate the GWAS summary data is available here: https://github.com/MRCIEU/BiobankGWAS/

Code used for the other statistical analysis in the study is available here: https://github.com/isadreev/UKBB_replication

## Author contributions

GH and TRG conceived, supervised and acquired funding for the study

IIS and GH performed the simulation study and empirical MR analyses

REM, GH, LP, PCH designed the UK Biobank QC and GWAS

LACM derived cleaned UK Biobank phenotype data using the PHESANT package

BLE performed the UK Biobank GWAS

IIS and GH wrote the first draft

All authors contributed to critical analysis of the results and commented on the paper

