## Supplementary Information for "Navigating sample overlap, winner’s curse and weak instrument bias in Mendelian randomization studies using the UK Biobank"

Sadreev et al

### Supplementary tables

Supplementary table 1: Meta-data for all 2514 traits analysed from the UK Biobank

Supplementary table 2: Results of independent GWAS significant associations discovered in the full UK Biobank analysis

### Supplementary figures

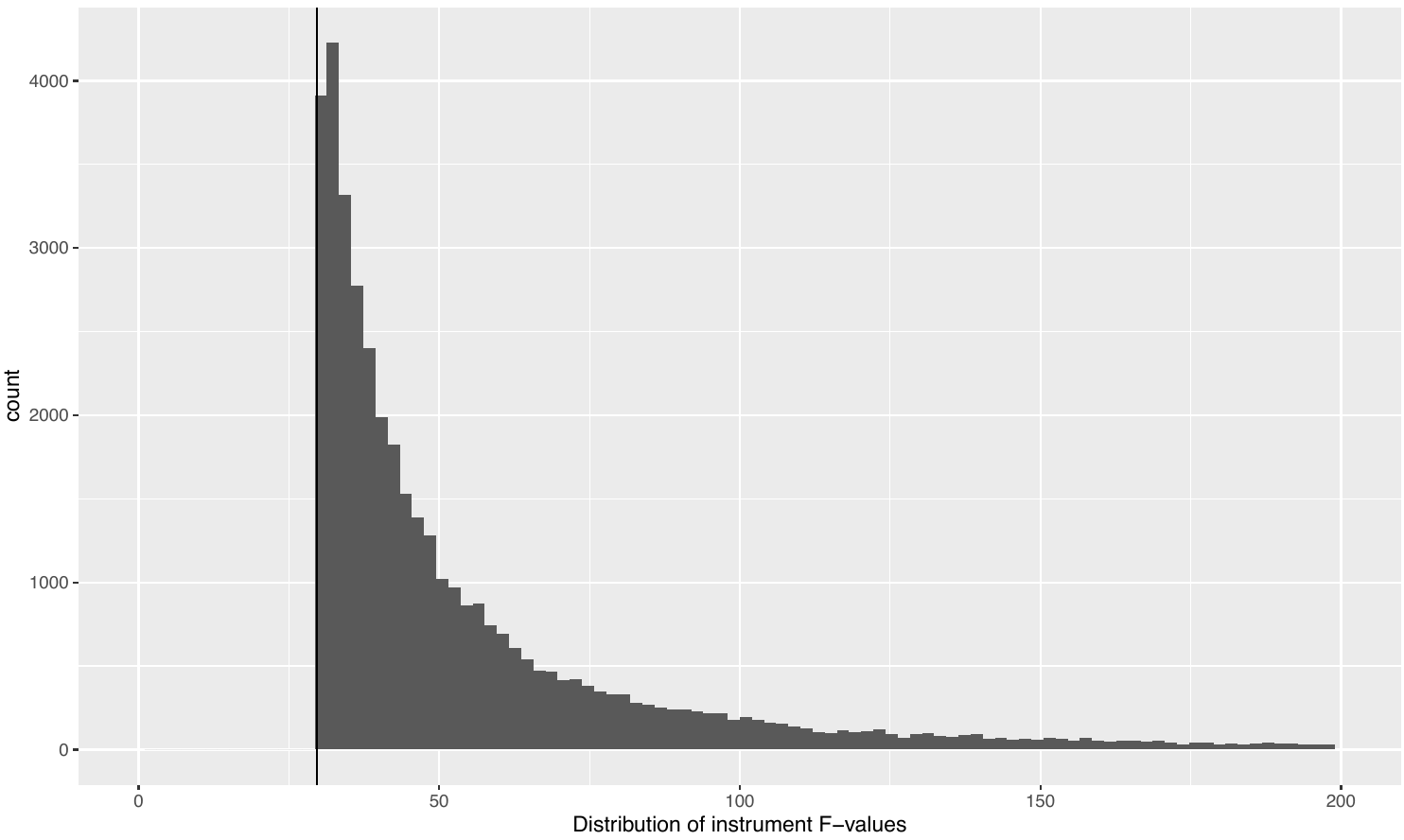

Supplementary figure 1. Distribution of F-statistics in the OpenGWAS summary database.

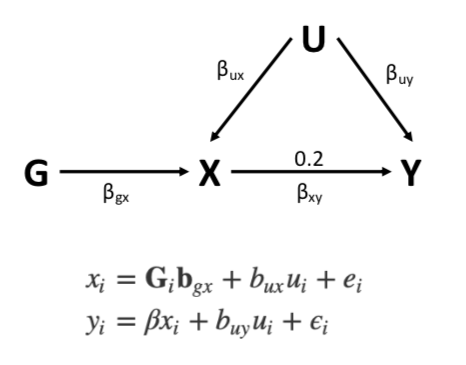

Supplementary figure 2. Scheme of the MR analysis. IVW was used to estimate SNP effects on x and y. The true value for the association was chosen to be 0.2 according to [Burgess et al 2016].

Supplementary figure 3. Simulations results for zero causal effect. A. Pure weak instrument bias. B. Replication used in different ways.

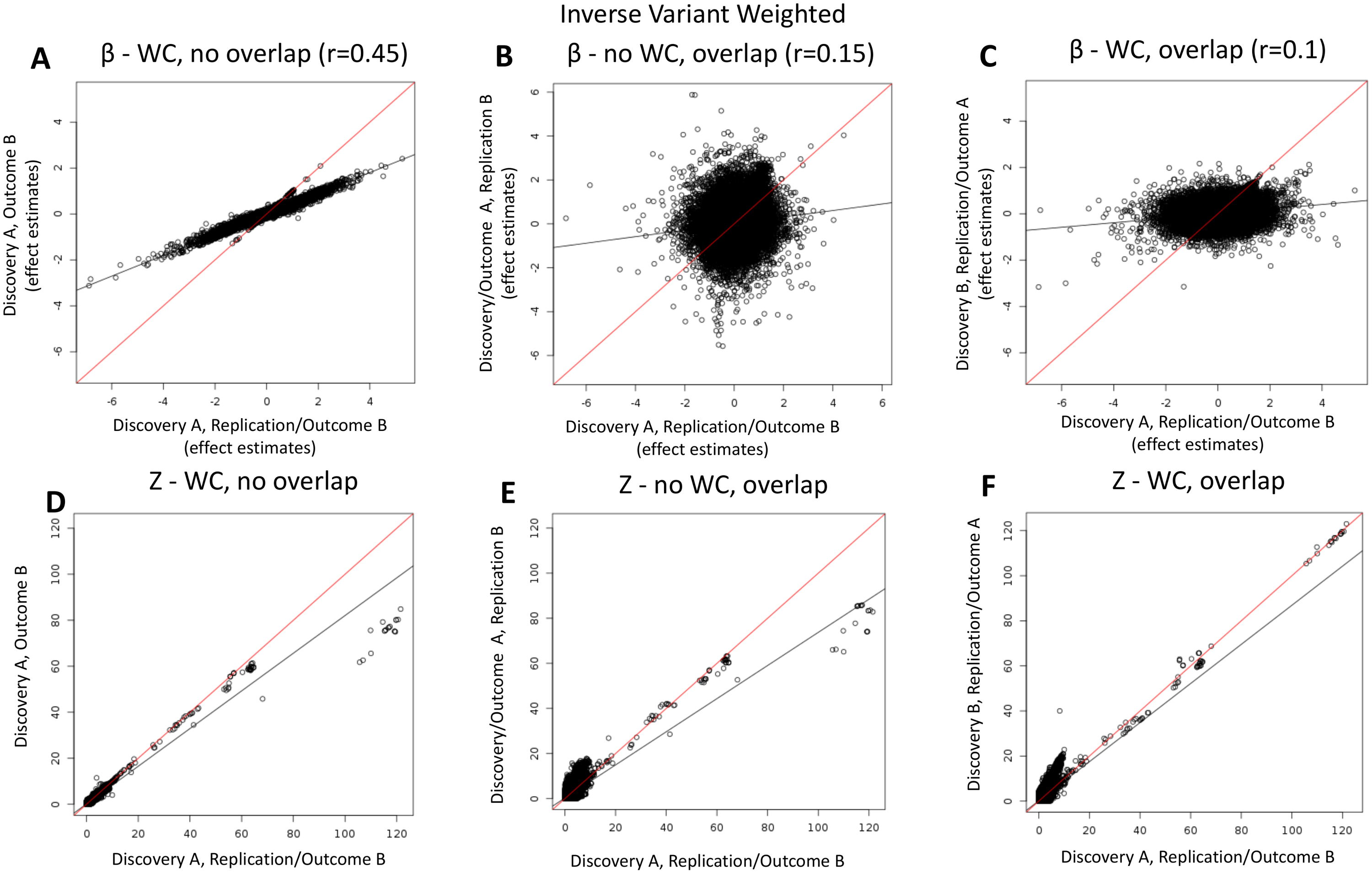

Supplementary figure 4. Effect estimates and Z scores for the case where all instruments are weak. X axis – always discovery from A, outcome and replication from B. A. Effect estimates for Winner’s curse and no overlap between discovery and outcome. B. Effect estimates for Winner’s curse and full overlap between discovery and outcome. C. Z scores for Winner’s curse and no overlap between discovery and outcome. D. Z scores for Winner’s curse and full overlap between discovery and outcome.
